# A Randomized Controlled Trial of N-Acetylcysteine in the Treatment of Early-Onset Preeclampsia: Study Protocol

**DOI:** 10.64898/2026.04.07.26350375

**Authors:** Kehinde S. Okunade, Adebola A. Adejimi, Muisi A. Adenekan, Iyabo Y. Ademuyiwa, Hameed Adelabu, Fatimah M. Habeebu-Adeyemi, Adaiah P. Soibi-Harry, Ololade Onasanya, Ayomide I. Fayinto, Temitope V. Adekanye, Fatimah Adeboje-Jimoh, Oziegbe Oghide, Nosimot O. Davies, Packson Akhenamen, Festus O. Olowoselu, Babasola O. Okusanya

**Affiliations:** Department of Obstetrics & Gynaecology, College of Medicine, University of Lagos, Lagos, Nigeria; Department of Obstetrics & Gynaecology, Lagos University Teaching Hospital, Lagos, Nigeria; Centre for Clinical Trials, Research, and Implementation Science, College of Medicine, University of Lagos, Surulere, Lagos, Nigeria; Department of Community Health & Primary Care, College of Medicine, University of Lagos, Lagos, Nigeria; Department of Obstetrics & Gynaecology, Lagos Island Maternity Hospital, Lagos, Nigeria; Department of Nursing Sciences, College of Medicine, University of Lagos, Lagos, Nigeria; Division of Infectious Diseases, Emory University School of Medicine, Atlanta, Georgia, United States; Accident and Emergency Department, Lagos University Teaching Hospital, Lagos, Nigeria; Department of Haematology & Blood Transfusion, College of Medicine, University of Lagos, Lagos, Nigeria

**Keywords:** Antioxidant, Disease progression, NAC, Nigeria, Severe preeclampsia

## Abstract

**Background:** Despite significant advancements in obstetric care, the incidence of preeclampsia remains a substantial public health challenge, and effective strategies to prevent the disease progression remain limited, particularly in low-resource settings. N-acetylcysteine (NAC), an antioxidant and glutathione precursor, has demonstrated anti-inflammatory and vasodilatory effects, making it a promising candidate for repurposing. However, robust evidence from well-powered randomized controlled trials is lacking.

**Objective:** This study will evaluate the impact of NAC on the time-to-disease progression in pregnant women with early-onset preeclampsia in Lagos, Nigeria.

**Methods:** This is the study protocol for a proof-of-concept, double-blind, randomized, controlled trial to be conducted between April 2026 to July 2028 at the maternity units of the two teaching hospitals in Lagos, Nigeria. At baseline, n=153 sexually active women aged 18 years or older diagnosed with early-onset preeclampsia at 24 to 34 weeks’ gestation will be randomised to receive either daily oral tablet containing 600 mg of NAC or a placebo tablet that is matched for appearance and the dosing regimen in addition to standard antenatal care from diagnosis (randomisation) until either 34 weeks’ gestation or delivery, whichever comes first. The primary endpoint is the time-to-progression (in days) of early-onset preeclampsia to severe disease. The data analysis will be conducted on an intention-to-treat basis. Kaplan-Meier estimates with a Log-rank test will be used to calculate and compare the time-to-disease progression for the treatment groups, while Cox proportional hazard models with a backwards conditional method will be used to compare the primary endpoint between the treatment arms while adjusting for other covariates for precision using hazard ratios (HRs) and 95% confidence intervals (95%CIs). Subgroup analyses will also be performed to assess the differential effects of significant covariates on the impact of NAC on disease progression. Statistical significance will be reported as *P*<0.05.

**Discussion:** This study will evaluate the efficacy of daily oral NAC compared to placebo in treating pregnant women with early-onset preeclampsia. If proven effective, NAC could offer a safe, affordable, and scalable intervention to reduce the burden of preeclampsia, particularly in resource-constrained settings.

**Registration:** PACTR202603712049354 (24th March 2026); https://pactr.samrc.ac.za/TrialDisplay.aspx?TrialID=3571

**Author Summary:** Preeclampsia is a serious complication of pregnancy that can lead to life-threatening outcomes for both mothers and babies, especially in low-resource settings. Although improvements in obstetric care have been made, there are still limited options to prevent the progression of early disease to more severe forms. N-acetylcysteine (NAC), a widely available and inexpensive medication with antioxidant and anti-inflammatory properties, has shown promise in early studies, but strong clinical evidence is lacking. In this study, we describe a randomized controlled trial designed to test whether NAC can slow the progression of early-onset preeclampsia in pregnant women in Lagos, Nigeria. Participants will receive either NAC or a placebo alongside standard care, and we will measure how long it takes for the disease to worsen. By using rigorous study methods, including blinding and intention-to-treat analysis, we aim to generate reliable evidence on the effectiveness of NAC in this context. If NAC is found to be effective, it could provide a safe, affordable, and scalable treatment option to improve maternal and newborn outcomes, particularly in settings where healthcare resources are limited.

## Introduction

Preeclampsia is a progressive multisystemic complication of pregnancy characterized by hypertension and often proteinuria, typically occurring after 20 weeks of gestation [5]. It affects approximately 2–8% of pregnancies globally [6] and up to 16.7% among pregnant women in Nigeria [7]. Preeclampsia remains a leading cause of maternal and perinatal morbidity and mortality worldwide, particularly in low- and middle-income countries [8]. Preeclampsia with severe features represents an escalation of preeclampsia severity and is associated with a higher risk of adverse maternal and fetal outcomes [6,9]. Severe features of preeclampsia include a systolic blood pressure of at least 160 mmHg or a diastolic blood pressure of at least 110 mmHg, thrombocytopenia, impaired liver function, severe persistent right upper quadrant pain, progressive renal insufficiency, pulmonary oedema, and new-onset cerebral or visual disturbances [10].

Despite significant advancements in obstetric care, the incidence of preeclampsia remains a substantial public health challenge [11] and effective strategies to prevent the disease progression remain limited, particularly in low-resource settings. While certain interventions, such as low-dose aspirin prophylaxis [12] in high-risk women, have shown some efficacy in reducing the risk of preeclampsia, there remains a need for additional measures to prevent the development of severe disease in women with early-onset preeclampsia, particularly in resource-constrained settings where the burden of disease is greatest [8]. Currently, the only cure for preeclampsia is delivery of the fetus and placenta [11,13] however, this is commonly associated with iatrogenic preterm delivery. To prevent this complication and improve maternal and perinatal outcomes, research efforts are currently focused not only on the prevention of preeclampsia but also on its treatment and prevention of its severe features.

The pathophysiology of preeclampsia is complex and not completely understood, but it is widely recognized that oxidative stress and inflammation play crucial roles in its development and progression [14]. N-acetylcysteine (NAC), a precursor to the antioxidant glutathione, has shown promise in reducing oxidative stress and inflammation [15]. The potential benefits of NAC extend beyond its antioxidant properties; it has also demonstrated anti-inflammatory and vasodilatory effects, making it a potential therapeutic agent for conditions characterized by oxidative stress and endothelial dysfunction, such as preeclampsia [15]. Recent preclinical and preliminary clinical studies [1–4] have suggested that NAC could mitigate the oxidative stress associated with preeclampsia, thereby reducing the progression to severe disease. However, despite the established long-standing safety profile of NAC in both general and obstetric populations [16–21], especially as an antidote for acetaminophen poisoning and as a mucolytic agent. There is a significant gap in the literature regarding definitive evidence of its efficacy in reducing the progression of preeclampsia in well-powered, randomised, controlled trials among women with early-onset preeclamptic pregnancies.

Our proposed proof-of-concept randomized controlled trial aims to address this knowledge gap by providing more robust data on the potential benefits of NAC in this context, evaluating its impact on the time-to-disease progression in pregnant women with early-onset preeclampsia in Lagos, Nigeria. The central hypothesis of the study is that NAC can be repurposed as an effective therapy to prolong disease progression in early-onset preeclamptic pregnant women. If proven effective in this context, NAC could provide a low-risk, affordable, and accessible intervention to reduce the burden of preeclampsia, which remains a significant cause of maternal and perinatal morbidity and mortality, particularly in low-resource settings.

## Patients and Methods

### Study design and setting

This is a proof-of-concept, multicenter, randomized, double-blind, placebo-controlled, superiority trial that will be conducted in the maternity units of two hospitals in Lagos, Nigeria, between April 2026 and July 2028. The hospitals, Lagos University Teaching Hospital (LUTH) in Idi-Araba and Lagos Island Maternity Hospital (LIMH) on Lagos Island, are two of the foremost public health institutions in Lagos State, southwest Nigeria, offering care to a population of over 20 million inhabitants [22]. These hospitals have established obstetrics departments with well-equipped and functional maternal units and jointly managed up to 1,787 cases of preeclampsia/eclampsia in 2024.

### Study population and eligibility criteria

Potentially eligible women in the two study sites are those aged 18 years or older diagnosed with early-onset preeclampsia at 24 to 34 weeks’ gestation [23]. Preeclampsia is defined as new-onset hypertension, characterised by systolic blood pressure of ≥140 mmHg or diastolic blood pressure of ≥90mmHg, measured with an appropriately sized cuff at least 4–6 hours apart, and accompanied by proteinuria, defined as ≥300mg in a 24-hour urine collection or ≥100 mg/L (equivalent to ≥2+ on dipstick urinalysis) in at least two random urine samples taken 4–6 hours apart after 20 weeks of gestation [6] and with no evidence of end-organ dysfunction. The exclusion criteria are women unwilling to provide informed consent, those with known allergy or hypersensitivity to NAC or any of its components, current use of medications or supplements that could interfere with the effects of NAC, such as other antioxidants, nitro-glycerine, activated charcoal, and chloroquine, receipt of a long course of mineral supplements containing NAC during the 6 months before enrolment, bleeding disorders, bronchial asthma, history of alcohol or drug abuse, active liver disease, significant chronic renal impairment, or major fetal anomalies, and women with preeclampsia with severe features or eclampsia at the time of randomisation.

### Study outcomes

The primary outcome measure is the time-to-disease progression (in days) of women with early-onset preeclampsia. Preeclampsia with severe features is the presence of specific clinical features, which may include blood pressure readings consistently elevated to 160/110mmHg or higher; severe proteinuria of 3+ or a urine protein/creatinine ratio of 0.3 or higher; platelet counts less than 100,000/µL; elevated liver enzymes (AST/ALT) or evidence of hepatic dysfunction; serum creatinine level greater than 1.1mg/dL or doubling of baseline creatinine in the absence of other renal diseases; symptoms and signs of pulmonary oedema such as dyspnoea and crackles on lung auscultation; cerebral or visual disturbances such as headaches, visual disturbances, altered mental status, seizures (eclampsia), or other neurological symptoms; and evidence of placental dysfunction such as placental abruption, fetal growth restriction or oligohydramnios on ultrasound examination [6,9]. Secondary outcomes will include (1) safety and treatment tolerability, (2) maternal outcome (maternal mortality), and (3) composite neonatal outcomes, including low birth weight in term neonates (baby’s birth weight less than 2500g), IUGR, stillbirth, NICU admission, and perinatal and neonatal mortality. Women who develop preeclampsia with severe features will be managed as per the obstetric unit protocol and will be excluded from further participation in the study.

### Sample size calculation

The sample size was calculated assuming a 1:1 randomization ratio, a two-sided type I error rate (α) of 0.05, and 80% statistical power to detect a hazard ratio of 0.5 between the NAC and placebo groups, based on an estimated 33.3% baseline probability of disease progression among pregnant women with early-onset preeclampsia [24]. Allowing for an anticipated 15% loss to follow-up or missing outcome data, the total planned sample size is 153 participants, who will be randomly allocated in equal proportions to the two study arms (approximately 76–77 participants per group).

### Study procedures

The study team will screen and enrol n=153 consecutively consenting pregnant women diagnosed with early-onset preeclampsia during their routine antenatal care or at the maternity units of the participating hospitals. We will enrol approximately 40% of participants at LUTH (n = 61) and 60% at LIMH (n = 92), reflecting differences in patient volume and prior recruitment performance. Once the study intent and procedures are introduced and informed consent obtained, baseline sociodemographic, obstetric, dietary, and laboratory (haemoglobin, liver function tests, kidney function tests, glutathione, and homocysteine status) information will be collected using an electronic case report form (CRF) on REDCap. A baseline ultrasound scan assessment will be done and documented. Gestational age is based on the date of the participant’s last menstrual period, which will be obtained at the time of random assignment or calculated from the baseline ultrasound biometry. Body mass index (BMI) is obtained by dividing the actual maternal weight (calculated by subtracting the estimated fetal weight at enrolment using a normogram chart from the measured maternal weight) by the maternal height (in kg/m^2^).

### Randomization and allocation concealment

Participants will be randomly assigned using a 1:1 block random sequence of 4 to 6 blocks generated by the study statistician from Random Allocation software version 1.0 (May 2004) prepared in advance of the trial to receive either a daily oral tablet containing 600 mg of NAC or a placebo tablet that is matched for appearance and the dosing regimen in addition to standard antenatal care from diagnosis (randomisation) until either 34 weeks’ gestation or delivery, whichever comes first [Figure 1]. The active drug (NAC) and placebo tablets will be packed in identical, coded, transparent dispensing sachets by the onsite pharmacy technician. The technician will then store the coded randomisation list in a sealed, opaque envelope, which will be kept in a locked file cabinet at the study site until the end of the study. The 600 mg oral daily dosage of NAC was selected after a careful review of the literature on the effectiveness of NAC in exerting antioxidant, anti-inflammatory, and antiproliferative effects while maintaining a favourable safety profile [16–21]. Furthermore, a low dose of 600 mg daily aligns with widely available commercial formulations that offer better gastrointestinal tolerability, which is important for sustained adherence in our study population and will also simplify procurement and ensure consistent dosing during the study. All study participants, irrespective of allocation to the intervention or placebo arm, will receive standard-of-care management for early-onset preeclampsia in accordance with the institutional protocol and internationally accepted guidelines. The study intervention will only be administered as an adjunct to standard care, which will include corticosteroids for fetal lung maturation, antihypertensive treatment, magnesium sulphate for seizure prophylaxis (when indicated), close maternal and fetal monitoring, and delivery based on standard clinical indications.

**F1_Figure 1.**
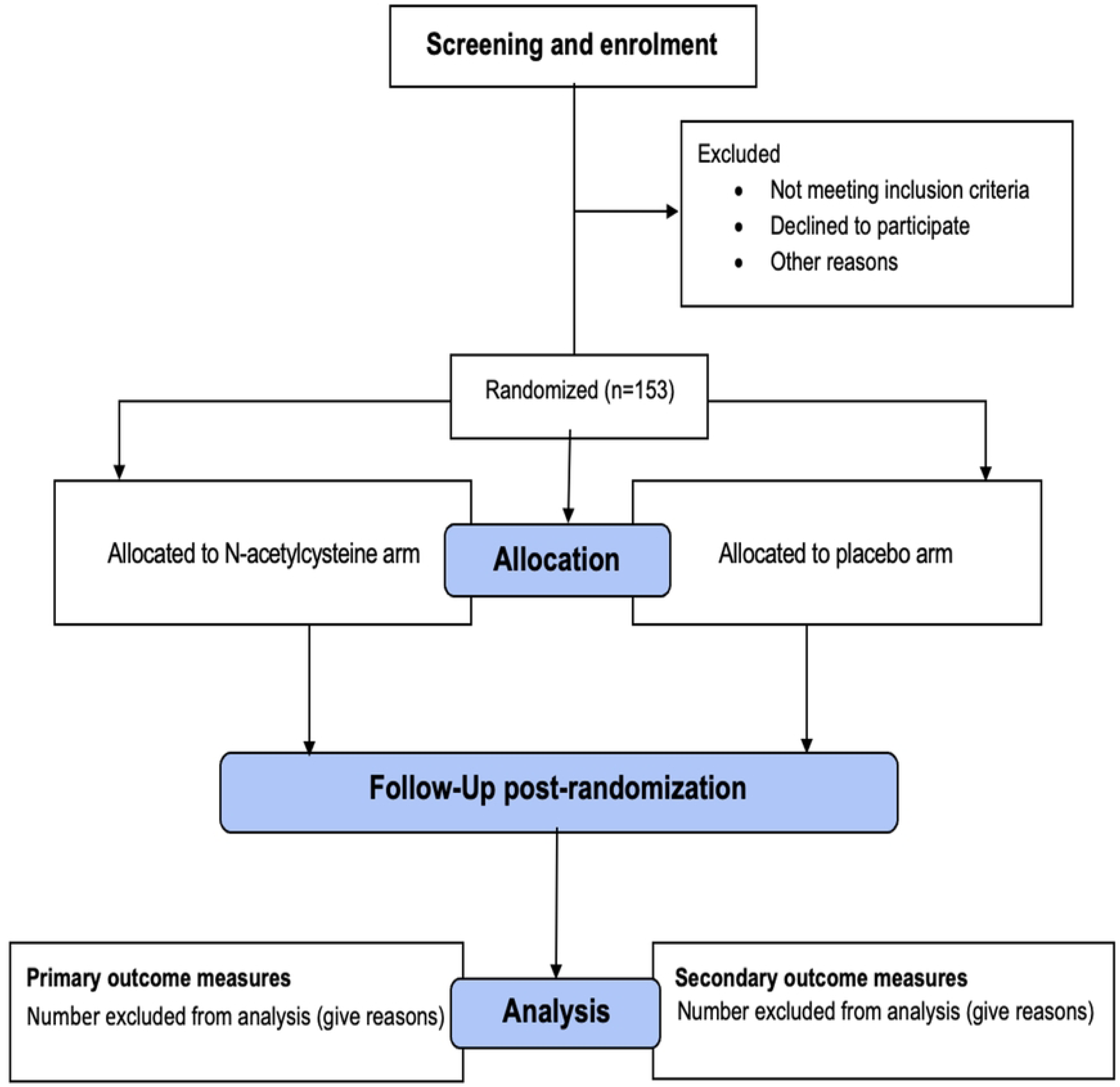
Trial flow chart

#### Follow-up evaluations

Data collection will commence on the day the participant is randomized into the trial and will continue until the trial is terminated or the participant withdraws from the trial at any time for any reason. Study visits will be scheduled every two weeks until 32 weeks and weekly thereafter until delivery. Participants will be monitored using a daily symptom diary to record features of severity—headaches, visual disturbances (e.g., blurred vision, scotoma), dyspnoea, and right hypochondria or epigastric pain and any adverse event—gastrointestinal upset, dry mouth, nausea, vomiting, diarrhoea, rash, and fatigue and its severity using a daily symptom diary and present this for recording at their next clinic visit. At every follow-up visit, a newly filled dispensing sachet containing the experimental drugs with the same numeric code will be given to each woman, and the pills remaining in used sachets are then counted to assess adherence. Adherence will be defined as the use of at least 70% of the prescribed tablets for each participant. Allocations to treatment arms will be concealed from the study investigators, clinic staff, and trial statistician using sealed opaque envelopes that are sequentially numbered. The onsite pharmacy technicians will not be involved in the design, conduct, statistical analysis, and reporting of study findings. Additional information to be collected during the follow-up visits includes a history of allergies, level of treatment adherence, drug tolerability, bedside urinalysis for albumin, venous blood samples for biweekly measurements of haemoglobin, creatinine, liver enzymes (AST/ALT), and complete blood count. Serum biochemical assessments of glutathione and homocysteine levels will be performed at randomisation (baseline) and at weeks 28 and 32. Fetal growth scans and umbilical artery velocimetry (from 32 weeks) will also be done biweekly until delivery. At delivery, data on gestational age (in weeks), total duration of experimental drug intake (in days), intrauterine growth restriction (IUGR), neonatal birth weight (in grams), neonatal status at birth, 7th- and 28th-day (dead/alive), neonatal intensive care unit (NICU) admission, and length of NICU admission (in days) will be collected and recorded in the electronic CRF. Cord blood sampling at delivery for analysis of glutathione and homocysteine levels will also be collected to provide additional insight into maternal-fetal antioxidant transfer and potential downstream effects of NAC.

### Management of adverse events

To ensure safety, participants will be educated on potential adverse events (AEs) such as gastrointestinal upset (nausea, vomiting, diarrhoea), rash, or rarely, allergic reactions and given 24/7 contact details of study staff. All AEs will be closely monitored at regular study visits using the symptom diaries to record any unusual symptoms or discomfort on each day, as well as between visits through scheduled check-in phone calls. Structured AE assessments will be conducted at each follow-up visit, and all AEs will be reported according to established standards [27]. Symptoms will be monitored without interruption of treatment unless bothersome to the participant. In the case of mild AEs, such as gastrointestinal upset, participants will be required to skip the next treatment dosage, and symptomatic treatment (e.g., antiemetics for nausea or vomiting) will be provided as appropriate. However, in case of persistent AE, the next treatment dosage will be delayed for 1 week and then completely discontinued if the AE persists. The PI may choose to discontinue a participant from the study for reasons such as serious AEs (SAEs), including significant renal or hepatic impairment or suspected unexpected serious adverse reaction (SUSAR), that is, a medical condition or situation in which continued participation in the study would not be in the best interest of the participant (codebreaking). Any SAE or SUSAR will trigger immediate clinical evaluation, and the event causing study discontinuation will then be documented in a dedicated CRF to capture the date and underlying reason for such discontinuation. Any participant experiencing an AE will be offered medical support, and our research insurance coverage will ensure that participants receive care at no cost as part of the study protocol. A Data and Safety Monitoring Committee (DSMC) will oversee safety evaluations throughout the study. The study will terminate if more than 30% of the enrolled patients experience SAE/SUSAR.

### Plan to improve treatment compliance

Strategies to improve treatment compliance include (1) comprehensive education sessions to inform patients about the importance of adherence to the treatment protocol and the potential benefits of the experimental treatment; (2) use of automated reminders through phone calls, text messages, or emails to remind patients of their upcoming clinic visits and treatment schedules; (3) offering transportation support or reimbursement for travel expenses to study sites; (4) regular monitoring and check-ins by research staff to address any concerns or adverse events promptly and to reinforce the importance of adherence; (5) provision of small incentives, such as gift cards or other tokens of appreciation, to encourage adherence to treatment protocol.

### Plan to secure trial drugs and quality assurance

The study experimental drug (N-acetylcysteine) is currently available and approved by the National Agency for Food and Drug Administration and Control (NAFDAC) as an antidote for acetaminophen (paracetamol) overdose, helping to prevent or mitigate liver damage. We will establish a preliminary agreement with a reputable pharmaceutical company with extensive experience providing clinical trial materials in Nigeria to supply the study drug (NAC). We will follow a stringent procurement process, including verifying the credibility and compliance of suppliers with regulatory standards and ensuring proper documentation and certification for each batch of trial drugs. To maintain the highest quality standards, we will ensure that the trial drug is (1.) manufactured in facilities that comply with Good Manufacturing Practice (GMP) guidelines and (2.) stored under optimal conditions as specified by the manufacturers. Regular audits of storage facilities and periodic random sampling and trial drug testing will be conducted throughout the study to ensure ongoing quality and consistency.

### Participants’ recruitment and retention strategies

These include (1) implementing robust community engagement strategies to review study protocol by a participants panel and advocacy group and provide suggestions for ensuring that the study and recruitment approach is acceptable and feasible for patients; (2) developing clear, concise, and engaging messages that highlight the importance of the study and its potential benefits to participants and society; (3) utilizing community organizations, patient advocacy groups, and healthcare providers to reach potential participants; (4) using other recruitment sites, if necessary, to screen and enrol participants into the trial; (5) ensuring that study staff are trained in cultural competence and can communicate effectively with participants from different backgrounds; (6) ensuring participants and study staff feedback to improve recruitment efforts continuously; (7) printing and posting of sensitization posters and fliers within and around the recruitment clinics; (8) completion of the participant’s locator form for each enrolled woman and review at each follow-up visit to ensure that the participant’s contact information is up to date; (9) using of automated contact and scheduling of follow-up through short text message reminders; (10) assigning study staff to monitor scheduling and ensure participants tracking through personal phone calls 48 hours before or after (for failure to show) their scheduled follow-up visits; and (11) reimbursement of transportation cost and time spent during study-related visit at five thousand naira (₦5,000), equivalent to three US Dollars ($3.00), for each visit.

### Quality control and data monitoring

All investigators and research assistants will be required to undergo training, including Good Clinical Practice (GCP) training, prior to the trial to ensure consistent practice. The training will cover a comprehensive understanding of the inclusion/exclusion criteria, follow-up protocols, and questionnaire completion. The research assistant will transfer identifiable data to an electronic database system housed in a secure facility at the trial site. Access to identifiable data will be limited solely to the principal investigator (KSO) throughout and after the trial concludes. The trial will undergo monitoring by quality assurance personnel from the research management office of the College of Medicine, University of Lagos, who will operate independently from the study team. Regular monitoring will occur to ensure the accuracy and quality of data throughout the study duration. Monitors will oversee and verify the essential documents (consent information, enrolment records, protocol deviations, and instances of loss to follow-up) for accuracy and completeness. An independent data and safety monitoring committee (DSMC) will also review the interim analyses to ensure the robustness of the data, treatment safety, and adherence to the trial protocol.

### Data management and statistical considerations

Data will be collected prospectively with continuous data entry and checking. Queries will be actively pursued to ensure prompt clarification. Data will be entered into the REDCap database and then exported to Stata version 18.0 for Windows (StataCorp LLC, Texas, USA) for statistical analyses. Missing or incomplete data will be handled by running a missing values analysis using pairwise or listwise deletion if no pattern is detected or the multiple imputation method if a pattern is detected [25]. The primary analysis will be conducted on an intention-to-treat basis. Kaplan-Meier estimates with a Log-rank test will be used to calculate and compare the time-to-disease progression for the treatment groups while considering deliveries without preeclampsia with severe features as censored observations. We will then perform Cox proportional hazard models with a backwards conditional method to compare the study outcomes between the treatment arms while adjusting for other covariates using hazard ratios (HRs) and 95% confidence intervals (95%CIs). We will assess the calibration or goodness-of-fit statistic of the final model using the Hosmer-Lemeshow test, while the model’s explanatory power will be reported with Nagelkerke’s pseudo-R². Subgroup analyses will also be performed to assess the differential effects of significant covariates on the impact of NAC on disease progression. With oversight from the DSMC, two interim analyses of efficacy and futility for the primary outcome (progression to severe disease) will be conducted at 50% and 75% enrolment using O’Brien-Fleming stopping criteria [26] at cutoff *p*-values of less than 0.013 and 0.025, respectively

### Ethical considerations

Approval for the trial was obtained from the research ethics committee of the Lagos University Teaching Hospital (LUTH-HREC – ADM/DSCST/HREC/APP/8031; February 27, 2026). The trial will be conducted in accordance with the Declaration of Helsinki. Participants will be informed about the study objectives, procedures, potential risks and benefits, and their right to withdraw at any time without any consequences to their care. Written informed consent will be obtained by the study investigators or trained clinical research assistants from all participants prior to enrolment. All data collected will be treated with strict confidentiality and used solely for this research. The trial and statistical analysis will be conducted with fidelity, and the reporting will adhere to the guidelines outlined in the Standard Protocol Items: Recommendations for Interventional Trials (SPIRIT) checklist. Clear and comprehensive standard operating procedures (SOPs) for participants’ treatment administration, data collection, follow-up evaluations, adverse event management and statistical analyses will be developed to align with the trial protocol. The study team will also conduct regular monitoring to ensure adherence to these SOPs. An independent data and safety monitoring committee (DSMC) will also review the interim analyses to ensure the robustness of the data, treatment safety, and adherence to the trial protocol.

### Community engagement plan

For our proposed trial, we plan to support comprehensive community engagement activities across the trial lifecycle: (1) Pre-trial engagement – We will identify and consult with key community stakeholders, including community-based organisations (CBOs), women’s health advocacy groups, and leaders of patient support networks; We will establish a community advisory board (CAB) composed of representatives from relevant CBOs, local health groups—Society of Gynaecology and Obstetrics of Nigeria (SOGON) and Nigerian Medical Association (NMA), and past patients to provide input on trial design, informed consent materials, and recruitment strategies; In partnership with participating sites and women’s groups, we will hold educational forums to introduce the study, its goals, and potential benefits, addressing myths and misconceptions about participation in clinical trials; (2) During the trial – We will organize regular feedback sessions with the CAB and participants to monitor perceptions, address concerns, and foster trust throughout trial implementation; We will work with trained community health workers to serve as liaisons for participants, helping navigate logistical barriers such as transportation or family-related hesitations that might hinder compliance or follow-up; We will develop and distribute translated and visually supported information materials to ensure comprehension and inclusiveness; (3) Post-trial engagement – Upon completion, we will share key findings with community members and stakeholders in accessible formats, including town hall meetings and infographic reports; We will engage with the community and local health authorities on how successful interventions or lessons can be adopted within ongoing health programs or advocacy campaigns.

**Table 1:** Time and Events Schedule.

| Study Phase | Screening | Treatment/Follow-up Phase (Gestational Age)<sup>a</sup> |  |  |  |  |  |  |  |  |  |  |  | End-of-Study |
| --- | --- | --- | --- | --- | --- | --- | --- | --- | --- | --- | --- | --- | --- | --- |
|  |  | 24 – 34 | 25 weeks | 26 weeks | 27 weeks | 28 weeks | 39 weeks | 30 weeks | 31 weeks | 32 weeks | 33 weeks | 34 weeks | Postpartum |  |
| Inclusion and exclusion criteria | X |  |  |  |  |  |  |  |  |  |  |  |  |  |
| Informed consent (ICF)<sup>c</sup> | X | X |  |  |  |  |  |  |  |  |  |  |  |  |
| Sociodemographic and clinical information | X | X |  |  |  |  |  |  |  |  |  |  |  |  |
| Randomization |  | X |  |  |  |  |  |  |  |  |  |  |  |  |
| Oral NAC/Placebo dispensing |  |  | X | X | X | X | X | X | X | X | X | X |  |  |
| Compliance assessment<sup>d</sup> |  |  | X | X | X | X | X | X | X | X | X | X |  |  |
| Follow-up evaluation visits |  |  |  | X | X | X | X | X | X | X | X | X | X |  |
| Primary endpoint (preeclampsia with severe features) |  |  |  |  |  | X | X | X | X | X | X | X |  |  |
| Secondary endpoints |  |  |  |  |  | X | X | X | X | X | X | X | X |  |
| Assessment of serum glutathione and homocysteine |  |  |  |  |  | X |  |  |  | X |  |  | X |  |
| CBC, Renal and Liver Function tests |  | X |  | X |  | X |  | X |  | X |  | X |  |  |
| Fetal growth scans and umbilical artery velocimetry |  |  |  |  |  |  |  |  |  | X |  | X |  |  |
| Postpartum outcomes (NICU admission, neonatal mortality) |  |  |  |  |  | X | X | X | X | X | X | X | X | X |
| Study close-out |  |  |  |  |  |  |  |  |  |  |  |  |  | X |
| Adverse event review<sup>e</sup> | Continuous review |  |  |  |  |  |  |  |  |  |  |  |  |  |

- a. There will be a minimum of 1 week between routine follow-up visits during pregnancy. - b. End of study is defined as 4 weeks postpartum or in case of early dropout. - c. Informed consent must be signed before any study-related procedures. - d. Drug compliance will be assessed via pill count and self-report thereafter. - e. Adverse events and concomitant medications will be monitored throughout the study, from the signing of the ICF onwards, until the participant's last study-related activity or until the participant has been deemed lost to follow-up after demonstrating due diligence in follow-up efforts.

## Discussion

Preeclampsia is a multisystem hypertensive disorder of pregnancy occurring after 20 weeks’ gestation [5] and affects about 2–8% of pregnancies globally [6], with a higher burden in Nigeria [7]. It remains a major cause of maternal and perinatal morbidity and mortality, particularly in low-resource settings [8]. Preeclampsia with severe features is associated with significant complications, including organ dysfunction and adverse fetal outcomes [6,9]. Despite advances in care, prevention of disease progression remains limited. Current strategies such as low-dose aspirin offer partial benefit, while delivery, the only definitive cure, often leads to preterm birth [12]. Therefore, there is a critical need for effective therapies that can delay progression to severe disease and improve outcomes. Oxidative stress and inflammation are key mechanisms in the pathogenesis of preeclampsia [14]. NAC, an antioxidant and glutathione precursor, has demonstrated anti-inflammatory and vasodilatory effects, making it a promising candidate for repurposing [15]. Preliminary studies suggest NAC may reduce oxidative stress and disease progression [1–4]; however, robust evidence from well-powered randomized controlled trials is lacking. This protocol, therefore, describes a proof-of-concept double-blind, randomized, controlled trial that will be conducted in the maternity units of two hospitals in Lagos, Nigeria, between April 2026 and July 2028 to evaluate whether NAC can delay progression to preeclampsia with severe features in women with early-onset preeclampsia. We have powered the study to detect the primary outcome that pregnant women aged 18 years or older with early-onset preeclampsia who had treatment with NAC will achieve delayed progression to severe disease. Therefore, if effective, NAC could offer a safe, affordable, and scalable intervention to reduce the burden of preeclampsia, particularly in resource-constrained settings.

### Dissemination of information

The trial will adhere to the reporting guidelines outlined in the Consolidated Standards of Reporting Trials (CONSORT) checklist, and the findings will be published in a peer-reviewed scientific journal. Any significant modifications to the trial protocol will be promptly communicated to the study investigators, LUTH-HREC, trial participants, funding agency, and the trial registry (PACTR).

### Trial Status

This manuscript details protocol version 3.0 dated March 25th, 2026. Recruitment for the trial has not yet begun as of the point of manuscript submission. Enrolment of participants is scheduled to commence in April 2026, with the final participant expected to be included in the trial by December 2027. The anticipated completion date for the trial is July 2028.

## Data Availability

No datasets were generated or analysed during the current study. All relevant data from this study will be made available upon study completion.

## Acknowledgement

The authors are particularly grateful to the personnel of the College of Medicine, University of Lagos’ Research Management Office (CMUL RMO) for their support in securing the funding for this study.

## Declarations

### Author contributions

KSO and AAA made substantial contributions to the concept or design of the work in this paper. KSO and AIF drafted the article. All the authors (KSO, AAA, MAA, IYA, HA, FMH, APS, OO, AIF, IYA, TVA, FA, OGH, NOD, PA, FOO, and BOO) revised the manuscript critically for important intellectual content and approved the version to be published.

### Availability of data and materials

No data are associated with this article. The authors intend to grant public access to the full protocol, participant-level dataset, and statistical code associated with this study.

## Supporting information caption

S1_Appendix – Approved Study Protocol Ethics

S2_Appendix – IRB approval

S3_Appendix – Clinical Trial Registration

S4_Appendix – SPIRIT checklist

S5_Appendix – Data Collection Form

